# Microstructural Brain Changes in Buerger’s Disease and Smokers: A Case-Control Study Using Diffusion Tensor Imaging

**DOI:** 10.1101/2024.11.15.24317118

**Authors:** Ali Asghar Asadollahi Shahir, Mohammad Hadi Gharib, Maryam Shahali Ramsheh, Reza Zahedpasha, Asma Razman, Abdollah Omidi, Pezhman Kharazm, Amir Ghaderi, Somayeh Ghorbani, Shervin-sadat Hashemian

## Abstract

**Aim:** Thromboangiitis Obliterans (TAO), or Buerger’s disease, affects peripheral vessels and is linked to smoking. This Diffusion tensor imaging tractography (DTI) study examines brain function changes in TAO patients compared to healthy smokers and non-smokers, aiming to reveal neurological implications beyond the known peripheral effects.

**Methods:** The case-control study involved 50 participants aged 27-56 in northeast Iran, including TAO patients, healthy smokers, and non-smoking controls. MRI scans with DTI were conducted to assess 21 brain tracts for fractional anisotropy (FA) and apparent diffusion coefficient (ADC).

**Results:** Significant differences in brain tract integrity were observed among the groups. TAO patients showed lower FA values in the minor forceps compared to healthy smokers, while smokers had higher FA values than non-smoking controls. ADC values were notably higher in TAO patients across several tracts, including corticospinal tracts, fornix tracts, arcuate fasciculus, and inferior longitudinal fasciculus, compared to both healthy smokers and controls.

**Conclusion:** The study highlights distinct brain tract alterations in TAO patients and suggests potential neurological consequences associated with the disease and smoking habits. DTI proves valuable in understanding microstructural brain changes and could serve as a diagnostic tool for evaluating smoking-related neurologic complications, providing insights into TAO’s impact beyond peripheral vessels.

**Key Points:**

1. Advanced MRI techniques, particularly DTI, reveal significant differences in fractional anisotropy (FA) and apparent diffusion coefficient (ADC) in the brains of individuals with Buerger’s disease and smokers.
2. Notable ADC variations in corticospinal and fornix tracts are observed between Buerger’s patients, smokers, and non-smokers.
3. FA values in the superior longitudinal fasciculus and the minor and major forceps significantly differ among Buerger’s patients, smokers, and controls, highlighting microstructural brain alterations.

## Introduction

Buerger’s disease, also known as thromboangiitis obliterans (TAO), is a vascular inflammatory condition primarily affecting the smaller and medium-sized blood vessels in the peripheral extremities. This disorder demonstrates a significant correlation with tobacco consumption, as noted in contemporary medical literature(1).

TAO prevalence exhibits marked global variability. Higher incidence rates are observed in regions ranging from the Mediterranean to Asia, including parts of Eastern Europe and South America. This distribution suggests a complex interplay of environmental, genetic, and socioeconomic factors influencing the disease’s occurrence(2).

Patients with TAO typically present with ischemic symptoms affecting distal arteries of the extremities, often manifesting as claudication in the foot arch and calf, and may include Raynaud phenomenon or livedo reticularis, causing pain in the hands, feet, and digits at rest(3). The etiology of Buerger’s disease is hypothesized to involve autoimmune mechanisms exacerbated by the effects of tobacco on the immune system(3).

Currently, there is no definitive diagnostic method for TAO, and the clinical criteria for diagnosing Buerger’s disease typically include a significant history of tobacco use, disease onset before the age of 50, occlusion of the infrapopliteal arteries, involvement of the upper limbs or presence of migratory thrombophlebitis, and the exclusion of other risk factors for atherosclerosis. (4).

All these elements are crucial for diagnosing thromboangiitis obliterans (TAO). While there is no definitive laboratory test for TAO, a comprehensive serological profile is essential to rule out other vasculitides that may present similarly. Acute phase reactants, such as C-reactive protein, which are typically elevated in other vasculitides, are not elevated in Buerger’s disease(4).

Digital subtraction angiography is considered the definitive diagnostic method for Buerger’s disease, despite the absence of specific angiographic features that are unique to the condition. The condition typically impacts the peripheral circulation, with primary involvement in the infrapopliteal region of the lower limbs and the areas beyond the brachial artery in the upper limbs(5).

Establishing the relationship between smoking and brain structure presents considerable challenges due to the intersection of various cardiovascular, psychiatric, and demographic risk factors that influence brain morphometry. Smoking is frequently linked with other substance use disorders, particularly heavy alcohol consumption. The adverse effects of smoking, which include oxidative stress, inflammation, and atherosclerosis, can potentially result in brain atrophy. A meta-analysis encompassing 761 individuals who smoke indicated that smoking is correlated with reduced volumes in several brain regions, including the mediodorsal thalamus, right cerebellum, left parahippocampus, left insula, and various prefrontal cortical areas(6).

Structural MRI research has revealed significant brain alterations associated with chronic smoking. Structural MRI studies suggest gray matter reductions across multiple cortical and subcortical regions. Diffusion tensor imaging has identified changes in white matter integrity, particularly in frontal areas and major fiber tracts. Functional imaging studies indicate altered connectivity patterns and resting-state activity in smokers. However, many of these findings come from studies with limited sample sizes, necessitating further research with larger cohorts to confirm these observations.(7).

Neuroimaging research on tobacco dependence reveals that it is associated with structural alterations in the brain, particularly within the white matter. These alterations are intricately linked to the mechanisms underlying nicotine addiction. Currently, diffusion tensor imaging (DTI) stands as the sole non-invasive technique capable of depicting the microstructure, morphology, and density of white matter fibers. The FA value obtained through DTI is widely utilized to indicate the structural integrity of white matter(8).

Diffusion tensor imaging (DTI) leverages the diffusion anisotropy of water molecules to generate images, quantitatively assess their dispersion within tissues, and decompose and quantify diffusion anisotropy data from a three-dimensional perspective. This technique can elucidate detailed tissue microstructures, including the distribution of white matter, the trajectory of fiber bundles, and in vivo changes in the myelin sheath. The fractional anisotropy (FA) value, commonly employed in DTI, is sensitive to neuronal attributes such as axon density, size density, and myelin formation(8).

DTI analysis incorporates several coefficients derived from the corresponding diffusion tensor, with the most commonly utilized being mean diffusivity (MD), fractional anisotropy (FA), axial diffusivity (AD), apparent diffusion coefficient (ADC), and radial diffusivity (RD)(9).

The aim of this case-control study was to employ diffusion tensor imaging (DTI) to investigate microstructural brain differences between patients with Buerger’s disease, healthy smokers, and healthy non-smokers. Specifically, the study utilized fractional anisotropy (FA) and apparent diffusion coefficient (ADC) measures to assess potential alterations in white matter tract integrity and diffusion characteristics in Buerger’s disease patients compared to healthy smokers and non-smokers. This examination was conducted to elucidate the neurological implications of the disease beyond its known peripheral vascular effects.

## Methods

### Study design

This case-control study was conducted in a city in northeast Iran, involving 50 participants aged 27 to 56 years. Subjects were patients referred to a radiology department for MRI by a radiologist. The study protocol was approved by the ethics committee.

### Study procedure

Necessary permits were obtained from relevant authorities to commence the study. Participants were categorized into three distinct groups based on specific entry criteria: healthy controls, patients diagnosed with Buerger’s disease, and healthy smokers. Following interviews and the acquisition of informed consent, MRI scans were performed. Inclusion criteria required participants to have a definitive diagnosis of Buerger’s disease. Nicotine dependence was confirmed in healthy nicotine-dependent individuals using the Fagerström Test for Nicotine Dependence(10). Participants were included if they had not tried to quit smoking or had been abstinent for more than three months in the past year. For the healthy non-nicotine-dependent controls, non-dependence on nicotine was confirmed with a Fagerström score indicating no dependence.All participants provided informed consent and followed the examination schedule. Age-, education-, and gender-matched non-smoking controls were recruited. Control subjects had smoked fewer than five cigarettes lifetime and none in the past five years. To minimize second-hand smoke exposure, controls were selected from smoke-free dormitories and had non-smoking parents.Exclusion criteria included any non-cooperation of the patient at any stage of the research. Additionally, the presence of severe mental illnesses (comorbidities) and degenerative brain diseases such as dementia and Alzheimer’s were grounds for exclusion. Other substance addictions and contraindications for MRI, such as having a metal prosthesis in the body, were also considered exclusion criteria.

### Image acquisition techniques and DTI parameters

The most common indicator used in this study was FA, a standardized measure ranging from 0 to 1 that indicates the degree of anisotropic diffusion of water within the micro- and macro-structure. Axial diffusivity (AD) and radial diffusivity (RD) measure diffusion along directions parallel and perpendicular, respectively, to the principal axis of the diffusion tensor. An increase in AD suggests enhanced structural organization, whereas a rise in RD is indicative of reduced structural coherence(11).

### DTI Analysis

In this study, a comprehensive assessment of brain microstructure was conducted using diffusion tensor imaging (DTI) across 21 specific white matter tracts. For each tract, both FA and ADC were calculated to evaluate structural integrity and diffusion characteristics.

1. Left-uncinate fasciculus
2. Right-uncinate fasciculus
3. Major forceps
4. Minor forceps
5. Left Corticospinal tracts
6. Right Corticospinal tracts
7. Left FORNIX
8. Right FORNIX
9. Right arcuate fasciculus
10. Right inferior longitudinal fasciculus
11. Right superior longitudinal fasciculus
12. Left arcuate fasciculus
13. Left inferior longitudinal fasciculus
14. Left superior longitudinal fasciculus
15. Corpus callosum
16. Right anterior cingulate
17. Right intermediate cingulate
18. Right posterior cingulate
19. Left anterior cingulate
20. Left intermediate cingulate
21. Left posterior cingulate

### Statistical analysis

The normality of the data was assessed using the Shapiro-Wilk test and by examining histograms. Since the data were not normally distributed, comparisons between the two groups in MRI assessments were conducted using the Mann-Whitney U non-parametric test. All statistical tests were two-tailed, and a p-value of less than 0.05 was considered statistically significant. Given the rarity of Berger’s disease, the sample size for this group was limited. Despite the small sample size, appropriate statistical methods were employed, and the results are deemed scientifically valid.

## Results

A total of 50 individuals aged 27 to 56 years participated in the study. The mean age and standard deviation (SD) for Buerger’s disease (BD) patients were 39.4±11 years, while healthy smokers had a mean age of 41.5±7.7 years (see Table 1 for detailed demographic features).

**Table 1:** demographic features of study members

| variable |  | group |  |  | Test Statistics | P-value |
| --- | --- | --- | --- | --- | --- | --- |
|  |  | Healthy smokers | Berger patients | Healthy controls |  |  |
|  |  | N (%) or Median (Q1, Q3) | N (%) or Median (Q1, Q3) | N (%) or Median (Q1, Q3) |  |  |
| sex | Male | 9 (90) | 9 (90) | 13 (43.3) | - | - |
|  | Female | 1 (10) | 1 (10) | 17 (56.7) |  |  |
| ethnicity | Fars | 8 (80) | 8 (80) | 6 (20) |  |  |
|  | Turkman | 0 (0) | 1 (10) | 9 (30) | - | - |
|  | Sistani | 2 (20) | 0 (0) | 14 (46.7) |  |  |
|  | Other | 0 (0) | 1 (10) | 1 (3.3) |  |  |
| Age(years) |  | 43 (35, 48) | 51.5 (41, 65) | 39.5 (33, 47) | 17.07 | < 0.001 |

Various brain tracts were investigated, revealing numerous statistically significant differences. These significant differences are summarized in Table 2. Additionally, **Figure 1A-I** provides a graphical representation of the data, offering a clearer visualization of the substantial variations observed across different variables. This comprehensive analysis underscores the pronounced differences detected in the brain tracts among the study groups. The remaining tracts, where no significant relationships between variables were found, are included in the Supplementary Table.

**Figure 1:**
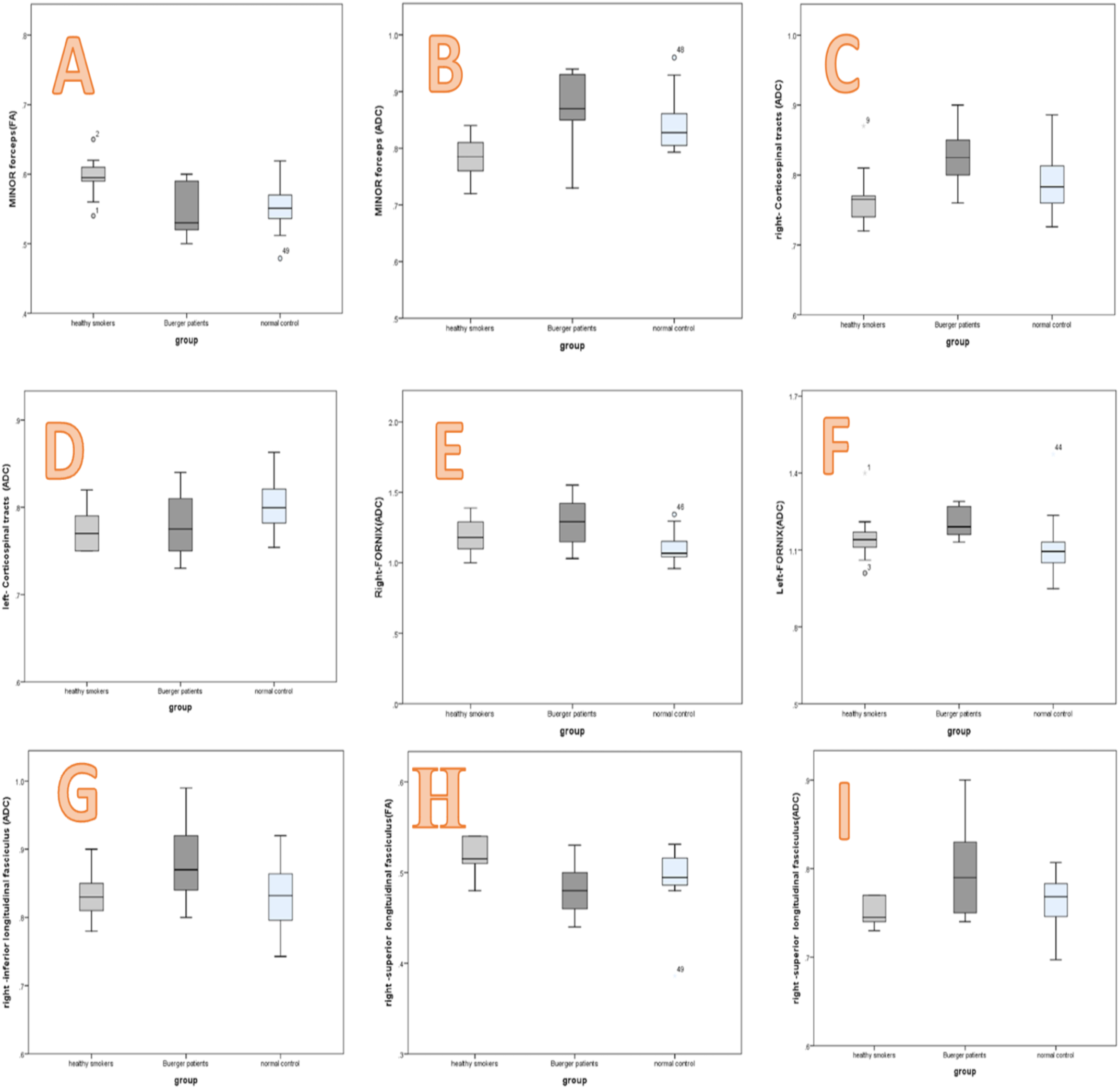
The left chart represents the healthy smoker group, the middle chart depicts the Buerger’s disease group, and the right chart illustrates the healthy control group. A: FA values of the minor forceps were lower in the Buerger’s disease (BD) group compared to healthy smokers, and higher in healthy smokers compared to controls. B: ADC values of the minor forceps were highest in the BD group, intermediate in smokers, and lowest in controls. C: Right corticospinal tract ADC values were higher in the BD group compared to both healthy smokers and controls. D: The left corticospinal tract ADC was significantly higher in healthy controls compared to healthy smokers. E & F: Higher ADC values were observed in the BD group for both the left and right fornix tracts compared to healthy controls. G: Increased ADC values in the BD group were found for the right inferior longitudinal fasciculus compared to both healthy smokers and controls. H: FA values for the right superior longitudinal fasciculus were lower in BD patients and higher in healthy smokers compared to controls. I: ADC values of the right superior longitudinal fasciculus were higher in the BD group compared to both healthy controls and smokers.

**Table 2:**
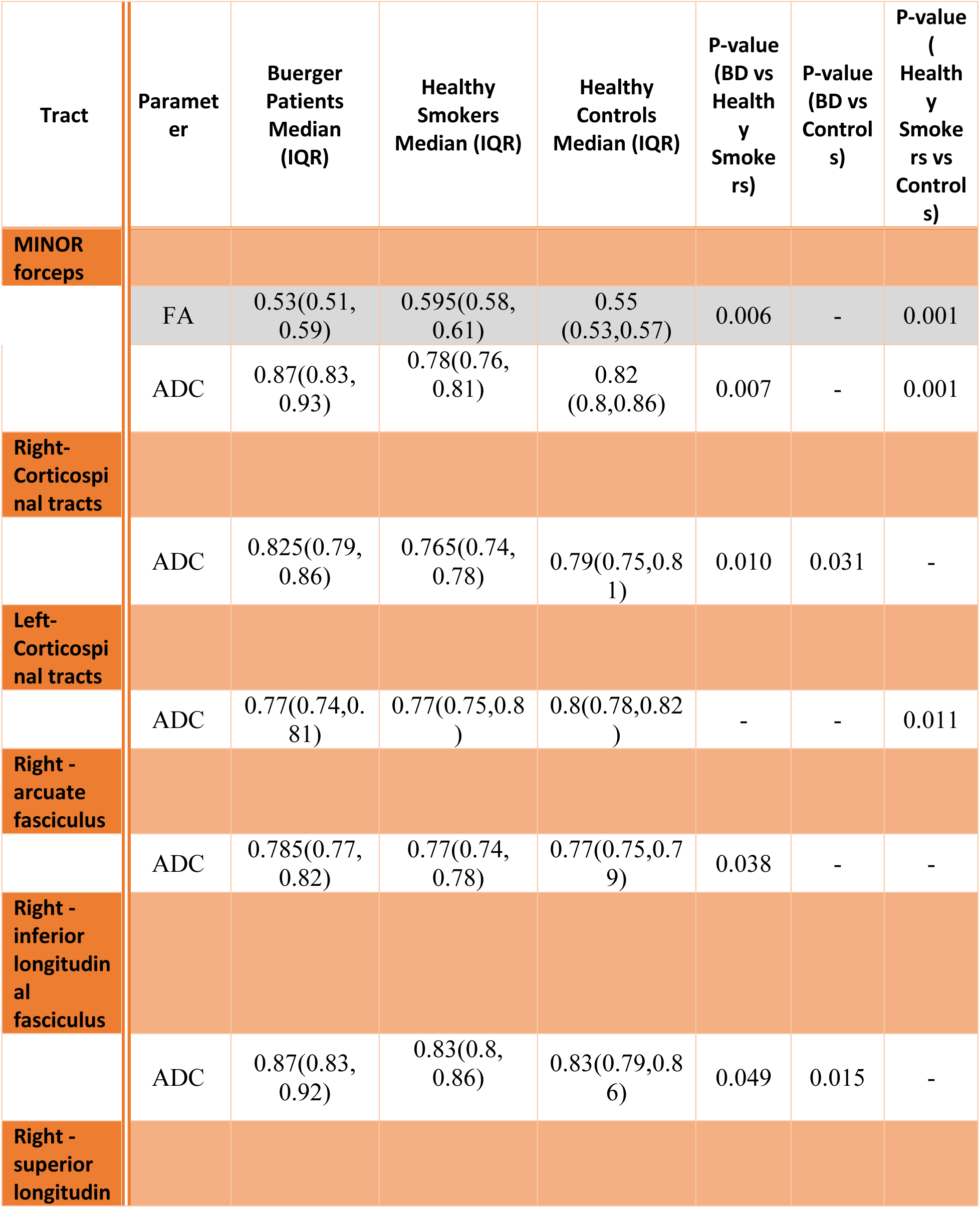
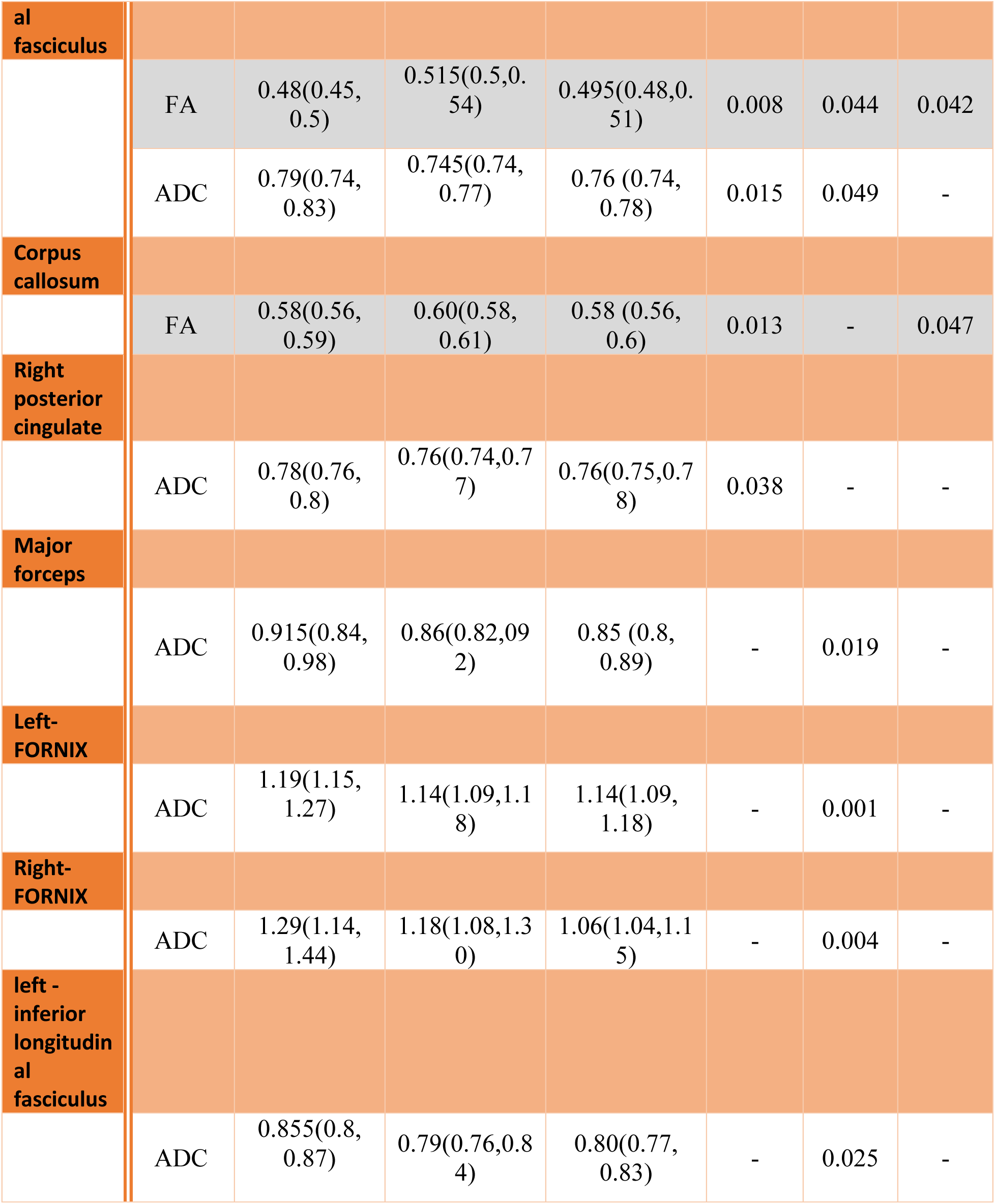
This table displays the tracts that were compared among all three groups and highlights the significant FA (Fractional Anisotropy) and ADC (Apparent Diffusion Coefficient) variables within these tracts. IQR= Interquartile Range.

To make the subject more tangible for readers, **Figure 2,3 A-C** illustrates the methods we employed and the process of data extraction. This figure depicts various brain tracts, with a table of fiber statistics displayed at the bottom of each image. These statistics correspond to the data extracted from each tract, which were subsequently subjected to statistical analysis.

**Figure 2.**
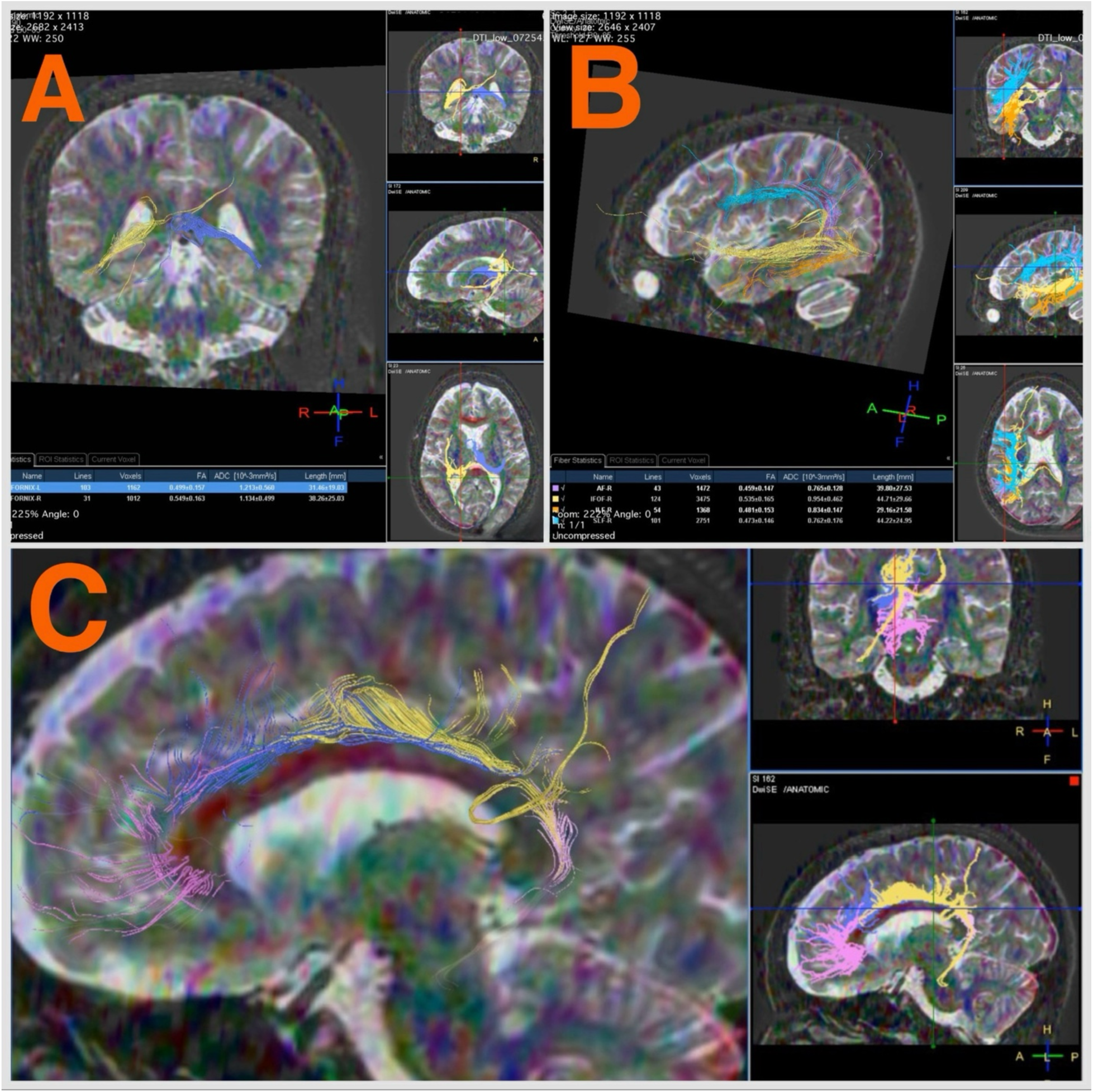
MRI photo of brain tracts, A: Fornix; B: superior longitudinal fasciculus, inferior longitudinal fasciculus, Inferior Fronto-occipital fasciculus; C: Cingulate.

**Figure 3.**
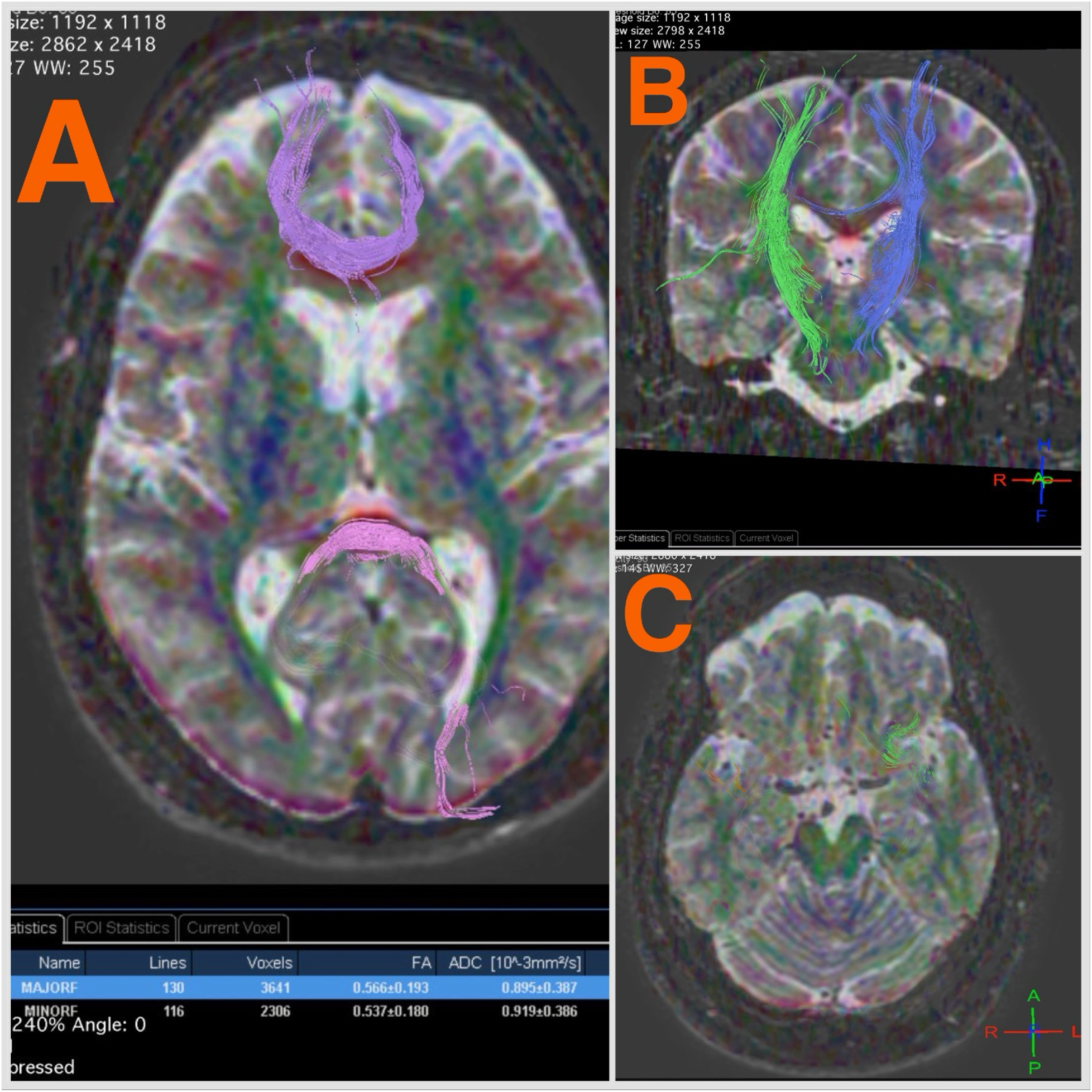
MRI photo of brain tracts, A: Anterior and posterior forceps; B: Cor=cospinal tracts; C: Uncinate fasciculus.

## DISCUSSION

The human brain is composed of highly localized motor and primary sensory regions, as well as functionally specialized areas dedicated to various cognitive processes. These regions often collaborate within a complex network known as a small-world network, essential for optimal brain function. Advanced network assessment aims to characterize networks using neurobiologically significant metrics derived from quantitative data sources like diffusion tensor imaging (DTI), functional magnetic resonance imaging (fMRI), electroencephalography (EEG), and magnetoencephalography (MEG)(12).

Among the crucial DTI coefficients, fractional anisotropy (FA) is particularly notable. Higher FA values within a tract indicate greater structural integrity. Conversely, FA tends to be lower in conditions like Alzheimer’s disease (AD)(13). Another key DTI metric, the apparent diffusion coefficient (ADC), reflects the rate of water diffusion in tissues: higher ADC values indicate increased diffusion rates, resulting in lower signal intensity on diffusion-weighted images, whereas lower ADC values produce higher observed signal intensity(14).

Cigarette smoking, a prevalent substance dependence globally, remains a leading cause of preventable health issues and mortality. Epidemiological and neuropsychological studies consistently show diverse cognitive and neurological impacts associated with smoking. Chronic smokers frequently show deficits in overall cognitive function and reduced capabilities in areas such as short-term memory, cognitive adaptability, spatial learning and memory, and speed of information processing.

Structural MRI studies further highlight the impact of smoking on brain integrity, revealing reduced gray matter integrity in various brain regions, including the prefrontal cortex (PFC), anterior cingulate cortex (ACC), insula, thalamus, and cerebellum. In our study, we observed significant differences in DTI metrics (FA and ADC) across several tracts, including the minor forceps, left corticospinal tract (LCST), right superior longitudinal fasciculus (RSLF), and corpus callosum (CC), between the healthy smoker group and the healthy non-smoker control group (HC), indicating structural damage caused by smoking.

The corpus callosum links the left and right hemispheres of the brain through roughly 200 million myelinated fibers. (15). The forceps minor links the frontal lobes, while the forceps major connects the occipital lobes (15).In the study by Yu et al. (2015), increased FA was observed in various brain regions, including the corpus callosum, internal and external capsules, and superior and posterior corona radiata in smokers compared to controls (16). This finding aligns with our observation of higher FA in healthy smokers compared to healthy non-smokers, suggesting that smoking may temporarily elevate FA levels. However, our study highlights that in the Buerger’s disease (BD) group, FA in the corpus callosum was significantly reduced compared to healthy smokers, indicating that Buerger’s disease exacerbates damage beyond what is observed with smoking alone. The increased FA reported by Yu et al. in smokers may reflect early-stage changes or compensatory mechanisms that do not fully capture the extent of damage seen in BD patients(16).

In contrast, van Ewijk et al. (2015) reported increased FA and decreased MD in smokers within white matter regions such as the basal ganglia, thalamus, and corpus callosum, but did not assess the relationship with FTND scores (17). Our findings suggest that while healthy smokers exhibited some increases in FA, the BD group showed a significant reduction in FA within the corpus callosum, highlighting the more severe and persistent damage observed in BD patients compared to healthy smokers.

The study by Fujin Lin and colleagues (18) corroborates our results by demonstrating reduced FA in heavy smokers, indicative of axonal damage and disrupted myelin integrity. Meanwhile, Hudkins et al.(19) observed elevated FA in the prefrontal white matter and corpus callosum among smokers, suggesting that FA might temporarily increase with initial smoking exposure. Our results, showing a significant reduction in FA in the BD group, underscore the progressive and lasting damage caused by Buerger’s disease compared to smoking alone.

In our study, we observed distinct patterns in FA and ADC values across various brain tracts. Specifically, FA values of the minor forceps (MF) were highest in the healthy control (HC) group and lowest in the BD group, whereas ADC values exhibited an inverse trend. These findings suggest significant damage to the MF tract, likely influenced by both smoking and BD. The MF tract comprises fibers extending laterally from the genu of the corpus callosum, linking regions of the dorsolateral prefrontal cortex (DLPFC), including portions of the middle and superior frontal gyri. This is consistent with findings from C. Gobbi et al., who reported more pronounced FA reductions in the MF tract among patients experiencing depression and fatigue, corroborating our results(20). Our study examined the minor and major forceps, along with the corpus callosum, in the context of smoking and Buerger’s disease, revealing their distinct structural impacts. Given their critical role in brain connectivity and significant alterations observed, it is plausible that smoking may impair these functions.

The corticospinal tract, a pivotal neural pathway connecting the cerebral cortex to the spinal cord, is essential for voluntary motor control and muscle movement. Originating from the frontoparietal cortices, it descends through the brainstem and decussates in the medulla before traveling down the spinal cord(21). Chronic nicotine use has been shown to increase corticospinal excitability, with studies indicating heightened corticospinal excitability during nicotine withdrawal and altered intracortical facilitation following acute nicotine administration(22,23).

In our study, we observed significant differences in ADC values of the left and right corticospinal tracts among the groups. The right corticospinal tract exhibited higher ADC values in the Buerger’s disease (BD) group compared to healthy smokers and controls, while the left corticospinal tract showed higher ADC values in healthy controls compared to smokers. Importantly, FA values for both tracts did not differ significantly among groups, indicating that ADC changes are more pronounced in BD compared to nicotine effects.

Additionally, elevated ADC values in the right corticospinal tract of the BD group compared to healthy smokers, and higher ADC values in the left corticospinal tract of healthy smokers compared to controls, suggest potential damage to these tracts due to both Buerger’s disease and smoking. These findings align with previous studies (24,25), which indicated increased axial diffusivity in the corticospinal tract segments in chronic pain patients, suggesting compromised tract integrity.

The superior longitudinal fasciculus (SLF) plays a crucial role in various cognitive and motor functions, as evidenced by multiple studies. Tetsuo Koyama et al. demonstrated that reduced FA in the SLF and corticospinal tract can predict motor and cognitive outcomes post-stroke. Their findings emphasized the role of the SLF in functions such as short-term memory, motor coordination, language processing, and visual perception. They also observed that degeneration of the left SLF is connected to aphasia, whereas impairment of the right SLF is related to hemispatial neglect(26). In another review, the SLF was identified as a key association fiber bundle connecting the frontal lobe with other ipsilateral hemisphere areas. Despite its importance, there is significant confusion and overlap in defining its structure across different studies, which necessitates a standardized nomenclature for better anatomical and functional correlation (27).Additionally, M. Hudkins et al. explored the white matter integrity in early adulthood smokers, revealing increased FA in the left SLF among other regions. Their study suggested that smoking induces microstructural changes in several white matter areas, with significant correlations between FA, radial diffusivity (RD), and smoking exposure measured in pack-years. These alterations indicate that smoking has a cumulative effect on white matter properties, potentially serving as biomarkers for nicotine dependence (28). In comparison, our study found significant differences in the FA values of the superior longitudinal fasciculus (SLF) among Buerger’s disease (BD) patients, healthy smokers, and healthy controls. The FA value was notably lower in the BD group compared to healthy smokers and controls. Additionally, the ADC of the right SLF was higher in the BD group. This suggests that both smoking and Buerger’s disease contribute to microstructural damage in the SLF, with more pronounced effects observed in BD patients. Our findings are consistent with Tetsuo Koyama’s study, which indicated that reduced FA in the SLF is associated with impaired motor and cognitive outcomes following stroke(26). In contrast, the study by M. Hudkins et al. found increased FA in smokers, which may reflect a temporary response to initial smoking exposure(28). However, our study shows that in BD patients, FA in the SLF is significantly reduced, indicating more severe and persistent damage compared to the changes observed by Hudkins et al. in smokers.

Moreover, our analysis revealed significantly higher ADC values in both the left and right fornix tracts in the Buerger’s disease (BD) group compared to healthy controls, suggesting potential damage to these pathways associated with the disease. This finding is consistent with the role of the fornix as a key output pathway from the hippocampus, crucial for cognitive and memory functions. Fitzsimmons et al. reported similar disruptions in white matter integrity, noting bilateral FA reductions in the fornix of patients with schizophrenia (29). Notably, our results showed no significant differences in fornix tract ADC values between the other groups, underscoring the distinctive impact of Buerger’s disease on these tracts.

The inferior longitudinal fasciculus (ILF) is a vital white matter pathway that links the occipital and temporal regions of the brain, playing an essential role in visual processing and complex cognitive functions. Research by Tetsuo Koyama et al. has demonstrated that the ILF comprises distinct subcomponents crucial for integrating visual information with anterior temporal activities, which are pivotal for memory and emotional processing(30). Additionally, Tzeng et al. reviewed the extensive involvement of the ILF in visual cognition and its disruption in various neuropsychological disorders, highlighting its role in processing and modulating visual cues(31).Our study found that ADC values in the right ILF were significantly higher in the Buerger’s disease (BD) group compared to healthy smokers and controls, indicating greater microstructural damage in BD. No significant differences in ADC were observed between smokers and non-smokers. Unlike Nicole A. T. et al.’s findings of reduced FA in the ILF due to nicotine use(32), our study did not detect significant differences in FA values for the ILF.

The arcuate fasciculus is a key white matter tract linking the frontal, parietal, and temporal lobes of the brain, playing a crucial role in language processing, especially phonological aspects in the left hemisphere, and visuospatial and semantic processing in the right hemisphere(33). The uncinate fasciculus, another significant pathway, links the lateral orbitofrontal cortex with the anterior temporal lobes, and is involved in episodic memory, language, and social-emotional processing(34).A study by Zhou et al. found that young smokers exhibited changes in diffusion indices in these tracts, with increased FA and AD and decreased RD in the left uncinate fasciculus, and increased AD and RD with decreased FA in the right arcuate fasciculus. These changes were associated with nicotine dependence, suggesting that smoking may induce microstructural alterations in these pathways(35).In our study, we observed significantly higher ADC values in the right arcuate fasciculus in the Buerger’s disease (BD) group compared to healthy smokers, indicating greater microstructural damage in BD patients. However, no significant differences in FA or ADC values were found in either the arcuate or uncinate fasciculus in both hemispheres across the groups. This suggests a distinct pattern of white matter alterations in BD compared to smoking-related changes reported by Zhou et al.(35).

Studies using voxel-based morphometry (VBM) have shown that smokers have reduced grey matter volume in the anterior cingulate cortex (ACC) compared to nonsmokers(36–38). MRI analyses also reveal decreased cortical thickness in the cingulate and insular cortices among smokers(39). These regions are key components of the brain’s salience network, which is involved in addiction and psychiatric disorders. Prolonged nicotine exposure, especially during sensitive developmental periods such as adolescence, may lead to cortical thinning and other neurotoxic effects.

Our study found a significant association specifically with the right posterior cingulate. However, no significant relationships were observed in the following areas: right anterior cingulate (FA and ADC), right intermediate cingulate (FA and ADC), right posterior cingulate (FA), left anterior cingulate (FA and ADC), left intermediate cingulate (FA and ADC), and left posterior cingulate (FA and ADC). In terms of diffusion metrics, the right posterior cingulate ADC was significantly higher in Buerger patients compared to healthy smokers (p = 0.038), but no significant differences were found between Buerger patients and healthy controls or between healthy smokers and controls. This suggests that while smoking may influence cortical thinning, the observed effects are specific to certain brain regions and do not fully account for the widespread cortical changes seen in previous studies.

## CONCLUSION

In conclusion, our study conducted using diffusion tensor imaging (DTI) revealed notable alterations in brain tracts among individuals with Buerger’s disease and healthy smokers. By systematically comparing brain function across Buerger’s patients, healthy non-smokers (control group), and healthy smokers, we identified significant variations in several key tracts: minor and major forceps, left and right corticospinal tracts, left and right fornix tracts, right arcuate fasciculus tract, right inferior longitudinal fasciculus, right superior longitudinal fasciculus, left inferior longitudinal fasciculus, corpus callosum, and right posterior cingulate tract. These differences, discerned through FA and ADC measurements, suggest promising avenues for further research aimed at assessing and correlating patient phenotypes with underlying neurological changes.

## Supporting information

Table 1: Comparison of the levels of MRI-related factors among the three participant groups. Kruskal-Wallis test results and median (interquartile ra

## Data Availability

All data produced in the present study are available upon reasonable request to the authors

## Acknowledgement

N/A

## Ethical Approval

This study was conducted in accordance with the ethical principles outlined in the Declaration of Helsinki. The ethics committee of Kashan University of Medical Sciences approved the protocol (Ethical Code: IR.KAUMS.MEDNT.REC.1397.049), ensuring that all procedures adhered to internationally recognized ethical standards for research involving human subjects.

## Author’s contributions

Ali Asghar Asadollahi Shahir: Conceptualization, Validation, Investigation, Funding acquisition.

Mohammad Hadi Gharib: Validation, Formal analysis, Data Curation, Writing - Review & Editing, Supervision.

Maryam Shahali Ramsheh: Validation, Formal analysis, Data Curation.

Reza Zahedpasha: Investigation, Resources, Writing - Original Draft, Writing - Review & Editing

Asma Razman: Resources, Writing - Original Draft.

Pezhman Kharazm: Resources

Abdollah Omidi: Project administration, Funding acquisition

Amir Ghaderi: Validation, Resources

Somayeh Ghorbani: Methodology, Formal analysis

Shervin-sadat Hashemian: Visualization, Methodology

**Patient consent statement:** N/A

**Clinical trial registration:** N/A

**Data availability statement:** N/A

## List of Supporting Information

Supplementary table - Comparison of the levels of MRI-related factors among the three participant groups. Kruskal-Wallis test results and median (interquartile range) variable for each group.

