## Supplementary material for "Microstructural Brain Changes in Buerger’s Disease and Smokers: A Case-Control Study Using Diffusion Tensor Imaging": Table 1: Comparison of the levels of MRI-related factors among the three participant groups. Kruskal-Wallis test results and median (interquartile ra

*Table 1: Comparison of the levels of MRI-related factors among the three participant groups. Kruskal-Wallis test results and median (interquartile range) variable for each group.*

| variable | Group | | | Kruskal Wallis Test statistics | P-value |
|---|---|---|---|---|---|
| | normal control (n=30) median (IQR) | Buerger patients (n=10) median (IQR) | healthy smokers (n=10) median (IQR) | | |
| left-uncinate fasciculus (FA) | 0.49 (0.46, 0.51) | 0.47 (0.46, 0.50) | 0.48 (0.47, 0.50) | 0.950 | 0.622 |
| left-uncinate fasciculus (ADC) | 0.87 (0.84, 0.9) | 0.899(0.873, 0.940) | 0.879(0.84, 0.91) | 2.312 | 0.315 |
| right -uncinate fasciculus (FA) | 0.48 (0.46, 0.51) | 0.470(0.450, 0.485) | 0.487(0.46, 0.52) | 4.462 | 0.107 |
| right -uncinate fasciculus (ADC) | 0.88 (0.84,0.91) | 0.916(0.885, 0.930) | 0.881(0.86,0.91) | 4.384 | 0.112 |
| Major forceps (FA) | 0.57(0.55, 0.59) | 0.569(0.52, 0.60) | 0.575(0.56, 0.61) | 2.649 | 0.266 |
| Major forceps (ADC) | 0.85 (0.8 ,0.89) | 0.922(0.84, 0.982) | 0.854(0.82, 0.92) | 5.620 | 0.060 |
| MINOR forceps (FA)* | 0.55 (0.53,0.57) | 0.54 (0.51, 0.590) | 0.595(0.58, 0.61) | 12.969 | 0.002 |
| MINOR forceps (ADC)* | 0.84 (0.8,0.86) | 0.866(0.83, 0.930) | 0.78(0.76, 0.81) | 12.964 | 0.002 |
| left- Corticospinal tracts (FA) | 0.57 (0.56,0.59) | 0.580(0.57, 0.6) | 0.58(0.57, 0.59) | 1.656 | 0.437 |

| | | | | | |
|---|---|---|---|---|---|
| left- Corticospinal tracts (ADC)* | 0.80(0.78, 0.82) | 0.782(0.74, 0.81) | 0.77(0.75, 0.80) | 6.728 | 0.035 |
| right- Corticospinal tracts (FA) | 0.58(0.56,0.59) | 0.579(0.55, 0.60) | 0.60(0.58, 0.62) | 3.702 | 0.157 |
| right- Corticospinal tracts (ADC)* | 0.79(0.75,0.81) | 0.826(0.79, 0.86) | 0.770(0.74, 0.78) | 8.101 | 0.017 |
| Left-FORNIX(FA) | 0.5(0.48, 0.52) | 0.492(0.46, 0.52) | 0.51 (0.49, 0.53) | 2.726 | 0.256 |
| Left-FORNIX(ADC)* | 1.112(1.05, 1.1) | 1.208(1.15, 1.27) | 1.15(1.09, 1.18) | 12.543 | 0.002 |
| Right-FORNIX(FA) | 0.5 (0.47,0.52) | 0.488(0.04, 0.49) | 0.49(0.48, 0.50) | 2.807 | 0.246 |
| Right-FORNIX(ADC)* | 1.104(1.04,1.1) | 1.293(1.14, 1.44) | 1.18(1.08, 1.30) | 10.171 | 0.006 |
| Right -arcuate fasciculus (FA) | 0.499(0.48, 0.21) | 0.487(0.45, 0.52) | 0.51(0.48, 0.53) | 1.594 | 0.451 |
| Right -arcuate fasciculus (ADC) | 0.7(0.75,0.79) | 0.808(0.77, 0.82) | 0.76(0.74, 0.78) | 4.671 | 0.097 |
| right -inferior longitudinal fasciculus (FA) | 0.50(0.47,0.52) | 0.491(0.45, 0.51) | 0.51(0.49, 0.53) | 1.610 | 0.447 |
| right -inferior longitudinal fasciculus (ADC)* | 0.83(0.79,0.86) | 0.882(0.83, 0.92) | 0.83(0.80, 0.86) | 6.357 | 0.042 |
| right -superior longitudinal fasciculus (FA)* | 0.498(0.48,0.51) | 0.481(0.45,0.50) | 0.51(0.50, 0.54) | 9.297 | 0.010 |
| right -superior longitudinal fasciculus (ADC)* | 0.76(0.74,0.78) | 0.797(0.74, 0.83) | 0.75(0.74, 0.77) | 6.747 | 0.034 |
| left -arcuate fasciculus (FA) | 0.50(0.48,0.52) | 0.493(0.46, 0.52) | 0.51(0.48, 0.53) | 1.091 | 0.579 |

| | | | | | |
|---|---|---|---|---|---|
| left -arcuate fasciculus(ADC) | 0.76 (0.72, 0.77) | 0.786(0.74, 0.82) | 0.761(0.74, 0.78 ) | 4.755 | 0.093 |
| left -inferior longituidinal fasciculus(FA) | 0.50 (0.49,0.52) | 0.50(0.47, 0.52) | 0.50 (0.49, 0.51 ) | 0.423 | 0.809 |
| left -inferior longituidinal fasciculus(ADC) | 0.80(0.79, 0.83) | 0.844(0.83, 0.92) | 0.81(0.76, 0.84 ) | 5.167 | 0.075 |
| left -superior longitude fasciculus(FA) | 0.50(0.49, 0.51) | 0.498(0.47,0.52) | 0.51(0.50, 0.54 ) | 2.045 | 0.360 |
| left -superior longituid fasciculus (ADC) | 0.74(0.72,0.75) | 0.751(0.72, 0.77) | 0.72(0.71, 0.74 ) | 3.240 | 0.198 |
| Corpus callosum (FA)* | 0.58(0.56, 0.6) | 0.576(0.56 , 0.59) | 0.60(0.58, 0.61 ) | 6.023 | 0.049 |
| Corpus callosum (ADC) | 0.99(0.94,1.04) | 1.040(0.98, 1.10) ) | 0.97(0.93, 1.02) | 3.492 | 0.174 |
| Right anterior cingulate(FA) | 0.49(0.5,0.53) | 0.505(0.47, 0.52) | 0.51(0.49, 0.54 ) | 3.059 | 0.217 |
| Right anterior cingulate (ADC) | 0.8(0.76,0.8) | 0.820(0.78, 0.86) | 0.81(0.79, 0.83 ) | 2.280 | 0.320 |
| Right intermediate cingulate(FA) | 0.53(0.50, 0.55) | 0.528(0.50, 0.55) | 0.531(0.52, 0.55 ) | 0.192 | 0.909 |
| Right intermediate cingulate (ADC) | 0.82(0.78,0.85) | 0.820(0.79, 0.83) | 0.81(0.76, 0.86 ) | 0.290 | 0.865 |
| Right posterior cingulate(FA) | 0.51(0.49,0.52) | 0.507(0.48 , 0.53) | 0.52(0.48, 0.54 ) | 0.821 | 0.663 |

| | | | | | |
|---|---|---|---|---|---|
| Right posterior cingulate(ADC) | 0.76(0.72,0.78) | 0.789(0.77,0.82) | 0.75(0.74, 0.77 ) | 4.179 | 0.124 |
| left anterior cingulate(FA) | 0.52(0.5,0.53) | 0.518( 0.51,0.54) | 0.53(0.51, 0.55 ) | 2.608 | 0.271 |
| left anterior cingulate(ADC) | 0.78(0.76, 0.8) | 0.804( 0.77,0.82) | 0.78 (0.75, 0.81 ) | 2.694 | 0.260 |
| left intermediate cingulate(FA) | 0.54(0.52,0.57) | 0.533( 0.51,0.54) | 0.54 (0.52, 0.56) | 1.807 | 0.405 |
| left intermediate cingulate (ADC) | 0.81(0.79,0.82) | 0.820( 0.75, 0.85) | 0.83(0.80, 0.87 ) | 2.038 | 0.361 |
| left posterior cingulate(FA) | 0.52(0.51,0.54) | 0.525( 0.50, 0.54) | 0.53(0.50, 0.55) | 0.414 | 0.813 |
| left posterior cingulate(ADC) | 0.77(0.75,0.79) | 0.786( 0.74, 0.81) | 0.76(0.73, 0.77 ) | 2.527 | 0.283 |
